# Beyond the Canonical Loci of Brugada Syndrome, Takotsubo Cardiomyopathy and Primary Pulmonary Arterial Hypertension

**DOI:** 10.64898/2026.07.20.26358466

**Authors:** Isabel Elliott, Hayley Coull, Christopher Aldous Oldnall

## Abstract

**Background:** The rare cardiovascular diseases of Brugada Syndrome, takotsubo Cardiomyopathy, and primary pulmonary arterial hypertension remain comparatively under-investigated despite their substantial morbidity and mortality. While common cardiovascular diseases have benefited from extensive genomic and therapeutic research, the molecular mechanisms underlying rarer cardiovascular conditions remain incompletely understood. This study aimed to investigate the genetic architecture and potential causal protein mediators of these rare cardiovascular diseases using integrated genomic and proteomic analyses.

**Methods:** Genome-wide association analyses were performed within the UK Biobank cohort for Brugada syndrome, takotsubo cardiomyopathy, and pulmonary arterial hypertension. Significant loci were identified following standard quality-control procedures and annotated using publicly available genomic databases. Protein–protein interaction networks were generated using STRING to investigate known and predicted biological relationships among identified genes and to explore potential shared disease mechanisms. Proteomic Mendelian randomisation analyses were subsequently conducted using protein quantitative trait loci (pQTLs) and Generalised Summary Mendelian Randomisation (GSMR) to evaluate whether genetically predicted circulating protein levels were associated with disease risk.

**Results:** Distinct genetic architectures were observed across phenotypes. Brugada syndrome demonstrated a concentrated association signal within the *THSD7B* locus alongside a variant in *TRPC4*. Takotsubo cardiomyopathy demonstrated associations involving *ANKRD31* and *PREX1*, while primary pulmonary arterial hypertension identified loci involving *ANO10* and *SHF*. Several significant variants could not be mapped to annotated genes, particularly within takotsubo cardiomyopathy, suggesting potential contributions from non-coding or regulatory genomic regions. Interaction network analyses identified biologically plausible relationships between identified loci and established cardiovascular pathways. Evaluation of circulating proteins as potential mediators of disease risk identified limited evidence for causal effects, although *LPA* emerged as a candidate protein associated with takotsubo cardiomyopathy and other ill-defined heart diseases.

**Conclusions:** These findings demonstrate substantial genetic heterogeneity across Brugada syndrome, primary pulmonary arterial hypertension, and takotsubo cardiomyopathy whilst highlighting shared biological themes involving electrophysiological regulation, calcium signalling, inflammation, and tissue remodelling. The integration of genomic association analyses, interaction-network approaches, and proteomic causal inference provides additional insight into the molecular pathways underlying these three cardiovascular diseases and may help prioritise targets for future functional investigation.

## 1 Introduction

Cardiovascular disease (CVD) remains the leading cause of death globally, responsible for an estimated 19.8 million deaths each year (WHO, 2025). As a result, major research investment and public health initiatives have historically focused on high-incidence, high-burden cardiovascular conditions such as myocardial infarction and stroke (Roth et al., 2020). This focus has led to important advances in prevention, diagnosis and treatment (National Institute for Health and Care Research, 2025; UK Parliament, 2025). However, it has also meant that rarer cardiovascular diseases have received comparatively limited attention, despite their substantial clinical impact. Rare cardiovascular conditions – including takotsubo cardiomyopathy, Brugada syndrome, and primary pulmonary arterial hypertension – each affect relatively small populations but collectively represent a meaningful and often under-recognised burden (Li et al., 2025; Rattanawong et al., 2025; H. Huang et al., 2025; Minhas & et al., 2015). These diseases are frequently associated with significant morbidity, reduced quality of life, and in some cases, sudden cardiac death. In several instances incidence appears to be increasing, but this may be driven by improved detection, demographic change, and evolving environmental or immunological exposures (Ghadri et al., 2018). Yet their underlying biological mechanisms remain incompletely understood.

Brugada Syndrome is a result of abnormality to the heart’s electrical system. Conduction changes, known as arrhythmias, in Brugada Syndrome are characterised by right precordial ST-segment elevation on ECG (Shukla, Basile, Van Name, & Ahmed, 2025), which can exacerbated or unmasked by things including fever and certain drugs. (Antzelevitch, 2006) This is a result of changes to ion channels in the heart, which can cause cells to fire signals too early or conduct pulses irregularly. (Borchard & Hafner, 2000) These changes in electrical signalling is what increases the risk of arrhythmias, which can lead to syncope or even sudden cardiac death in those with Brugada Syndrome. Therefore current management includes minimising trigger exposure and, for those who are at high risk, implantable cardioverter-defibrillators. (NHS, 2026).

In takotsubo Cardiomyopathy, the heart is seemingly oversensitive to hormones, such as catecholamines, released when the body is under sudden stress. This surge of hormones can lead to left ventricular “ballooning” - the pattern typically seen in takotsubo - which leads to dysfunction in the hearts pumping mechanism. (Pelliccia, Kaski, Crea, & Camici, 2017) This defect can cause symptoms such as chest pain, breathlessness, and low blood pressure (Mizusawa & Wilde, 2012), though after the hormone production settles, the heart is usually capable of recovering in most cases, returning back to its normal mechanism. (British Heart Foundation, 2026).

Primary pulmonary arterial hypertension results from narrowing of the blood vessels in the lungs. (Lan, Massam, Kulkarni, & Lang, 2018) This is due to over-constriction of the pulmonary vessels, eventually leading to the thickening of their walls or even the development of clots inside from the increased pressure. (Lai, Potoka, Champion, Mora, & Gladwin, 2014) Therefore, blood flow is decreased and the right ventricle of the heart must work harder to pump blood through the lungs, which can lead to symptoms such as shortness of breath and chest pain. (Hoendermis, 2011)

A major challenge in these conditions is heterogeneity. Pathogenesis may involve complex interactions between genetic susceptibility, inflammatory and immune processes, myocardial remodelling, vascular dysfunction, and electrical instability (Frak et al., 2022). Clinical presentation can be variable, overlapping with more common cardiovascular disorders, leading to under diagnosis or delayed diagnosis (Scafa-Udriste, Horodinschi, Babos, & Dinu, 2024; Great Ormond Street Hospital, 2015). Therapeutic options are often limited and are commonly extrapolated from treatments for more prevalent cardiovascular diseases rather than derived from disease-specific biological insight. Genome-wide association studies (GWAS) provide an opportunity to identify genetic loci associated with disease risk and to generate hypotheses regarding the biological pathways involved in disease development. However, translating genetic associations into mechanistic understanding remains challenging, particularly when associated variants lie within non-coding regions or map to genes with poorly characterised functions.

Integrating genetic findings with proteomic data can help bridge this gap. Proteins represent key functional components of biological pathways and may provide insight into the mechanisms linking genetic variation to disease. Mendelian randomisation can further aid interpretation by evaluating whether genetically predicted protein levels are consistent with a potential causal role in disease pathogenesis (Sun et al., 2018). In this study, we investigate the genetic architecture of rare cardiovascular diseases; takotsubo cardiomyopathy, Brugada syndrome, and primary pulmonary arterial hypertension, using genome-wide association analyses and explore the biological relevance of identified signals through complementary proteomic and Mendelian randomisation approaches. By combining these methodologies, we aim to improve understanding of the molecular pathways underlying these conditions and identify targets for future investigation.

## 2 Methods

This study investigated the genetic architecture of three rare cardiovascular diseases: Brugada syndrome, takotsubo cardiomyopathy, and primary pulmonary arterial hypertension. Genome-wide association studies (GWAS) were performed within the UK Biobank to identify disease-associated loci. Significant variants were subsequently annotated and explored using gene mapping and protein–protein interaction analyses. Proteomic Mendelian randomisation analyses were then conducted to evaluate whether genetically predicted circulating protein levels were associated with disease risk.

Genome-wide significance for all analyses was defined as *p <* 5 *×* 10*^−^*^8^. For proteomic Mendelian randomisation analyses, false discovery rate (FDR) correction was applied across tested protein–disease associations, with an adjusted threshold of *q <* 0.05 considered statistically significant. All analyses were conducted using R.

### 2.1 Genome-Wide Association Studies

Genome-wide association studies were performed for single-nucleotide polymorphisms (SNPs) within the UK Biobank cohort. Individuals were restricted to those of European genetic ancestry to minimise population stratification. Standard quality control procedures were applied, including exclusion of variants with low imputation quality and extreme allele frequency values. Association testing was conducted under an additive genetic model using logistic regression for case–control phenotypes. Models were adjusted for age, sex, genotyping array, and principal components of ancestry. Genome-wide significance was defined as *p <* 5 *×* 10*^−^*^8^. Variants with allele frequencies approaching fixation were excluded to avoid unstable effect estimates. Manhattan plots were created per phenotype.

### 2.2 Variant Annotation and Biological Interpretation

Genome-wide significant variants identified from GWAS analyses were annotated using the GeneCards human gene database (GeneCards, 2025). Variants were mapped to nearby or overlapping genes where possible, and associated gene functions, biological pathways, and known disease relationships were reviewed to aid biological interpretation. Variants that could not be confidently assigned to annotated genes were retained and classified as unmapped loci. For loci mapping to protein-coding genes, protein–protein interaction analyses were performed using STRING-DB. Interaction networks were generated to investigate known and predicted biological relationships between encoded proteins, incorporating evidence only from curated databases and experimentally validated interactions to improve quality of evidence. These analyses were used to explore potential shared pathways and mechanistic relationships among genes identified across the investigated cardiovascular phenotypes.

### 2.3 Proteomic Mendelian Randomisation

To investigate whether circulating proteins may contribute to disease risk, Mendelian randomisation analyses were performed using Generalised Summary Mendelian Randomisation (GSMR). Mendelian randomisation analyses rely on three instrumental variable assumptions: (1) the genetic variants are associated with the exposure (relevance assumption); (2) the genetic variants are independent of confounders of the exposure–outcome relationship (independence assumption); and (3) the genetic variants influence the outcome only through the exposure of interest and not through alternative biological pathways (exclusion restriction assumption). Figure. 1 illustrates how GSMR incorporates multiple SNPs. In this study, SNPs identified from published protein quantitative trait locus (pQTL) studies were used as instrumental variables for circulating protein levels. Independent variants associated with protein levels at genome-wide significance (*p <* 5*×*10*^−^*^8^) were selected as instrumental variables.

**Figure 1:**
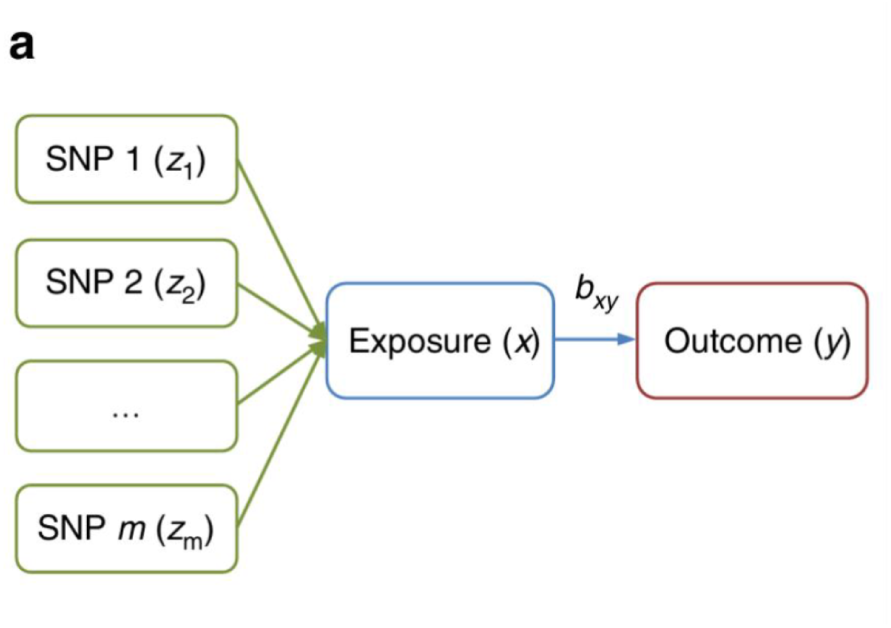
Schematic overview of the Mendelian randomisation framework used in this study. Independent genetic variants associated with circulating protein levels were used as instrumental variables to estimate the relationship between protein abundance and disease risk.

Linkage disequilibrium clumping was performed using an *r*^2^ threshold of 0.001 within a 10 Mb window to ensure instrument independence. Only cis-acting variants, defined as variants located within or proximal to the encoding gene, were retained to reduce the risk of horizontal pleiotropy. Effect alleles were harmonised between exposure and outcome datasets, and palindromic variants with ambiguous strand alignment were excluded. GSMR was used to estimate the association between genetically predicted protein levels and disease risk while accounting for residual linkage disequilibrium between variants. HEIDI outlier filtering was applied to identify and remove variants demonstrating evidence of horizontal pleiotropy.

## 3 Results

Genome-wide association analyses identified multiple significant loci across Brugada syndrome, and takotsubo cardiomyopathy and primary pulmonary arterial hypertension within the UK Biobank cohort. Distinct patterns of association were observed between phenotypes, with several loci mapping to genes involved in electrophysiological regulation, calcium signalling, cellular remodelling, inflammation, and endothelial dysfunction. In addition to established cardiovascular-related genes, a number of associations involved poorly characterised or non-coding loci, suggesting broader molecular heterogeneity across these three rare cardiovascular diseases. To further investigate potential biological relevance, identified loci were explored using protein–protein interaction analyses and downstream Mendelian randomisation approaches to prioritise candidate protein mediators of disease susceptibility.

### 3.1 Genome-wide significant loci identified across phenotypes with distinct interaction networks

Manhattan plots for each phenotype are presented in Figure 2. Genome-wide significant loci (*p <* 5 *×* 10*^−^*^8^) were identified across multiple phenotypes. Distinct patterns of genetic association were observed across analysed phenotypes, consistent with heterogeneous genetic architectures. 15 of the genome-wide significant variants, all associated with takotsubo cardiomyopathy, could not be mapped to annotated genes using the GeneCards database (Appendix A). Several significant variants were located within RNA genes corresponding to non-coding transcripts. These loci were retained for completeness but were not considered protein-coding candidates in downstream analyses.

**Figure 2:**
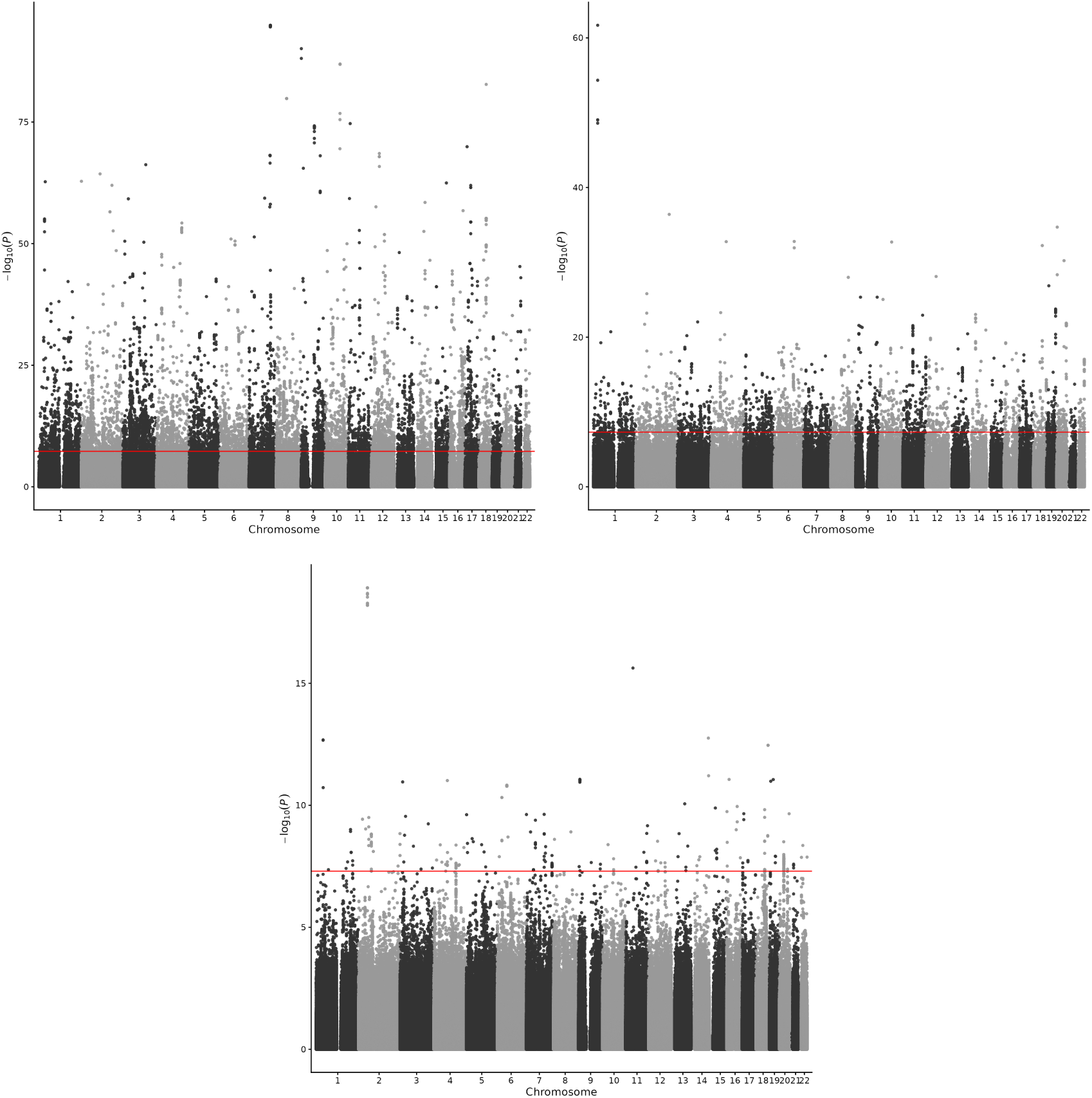
Manhattan plots showing genome-wide association results for rare cardiovascular diseases within the UK Biobank cohort. Reading from left-to-right; (A) Primary pulmonary arterial hypertension (ICD-10: I27.0), (B) Brugada syndrome (ICD-10 proxy: I49.8), and (C) Takotsubo cardiomyopathy (ICD-10 proxy: I51.8). The horizontal dashed line indicates the genome-wide significance threshold (*p <* 5 *×* 10*^−^*^8^).

Following functional annotation, STRING protein–protein interaction networks were generated to investigate known and predicted biological relationships between proteins encoded by loci identified in the genome-wide association analyses. Among the identified loci, *TRPC4* demonstrated interactions with proteins involved in developmental and signalling pathways, including *BMPR2*, a well-established pulmonary arterial hypertension susceptibility gene. Given the potential implications of this interaction for shared cardiovascular disease mechanisms, the *TRPC4* interaction network is presented in Figure. 3, while additional interaction networks are provided in Supplementary Figure 4.

**Figure 3:**
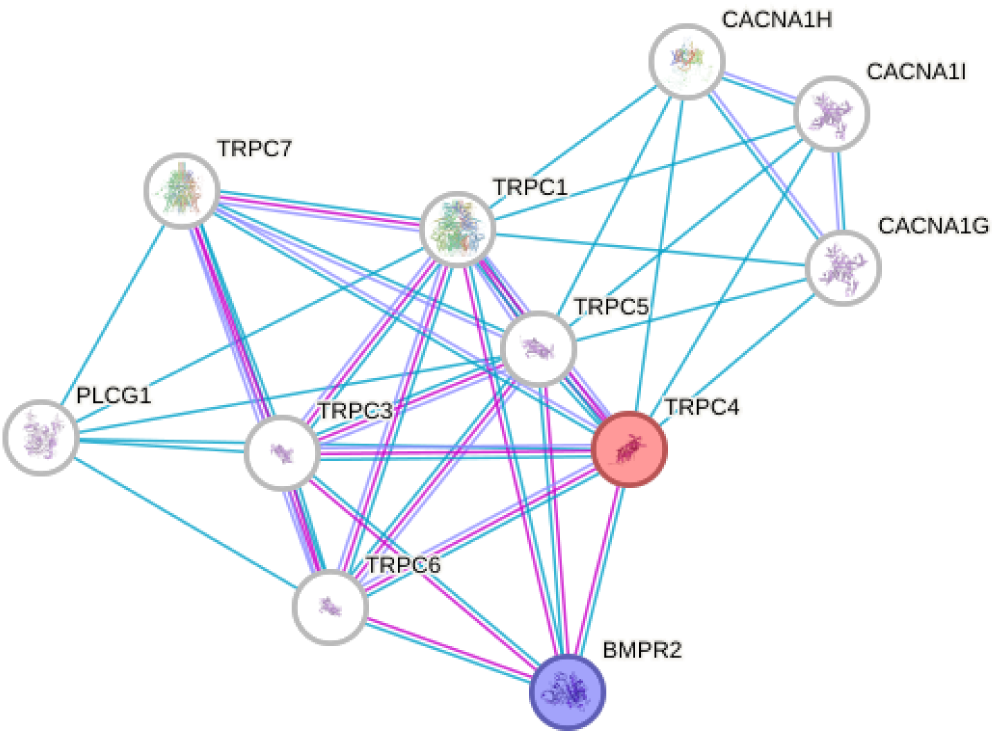
STRING protein–protein interaction network for *TRPC4*. Nodes represent proteins and edges represent known or predicted interactions derived from curated databases and experimental evidence. The observed interaction between TRPC4 and BMPR2 highlights a potential link between calcium-dependent signalling pathways and pulmonary arterial hypertension-associated biology.

#### Brugada Syndrome

For Brugada syndrome, seven genome-wide significant variants were identified. Six variants mapped to the *THSD7B* locus, indicating a concentrated association signal within this gene region. The remaining variant mapped to *TRPC4*. Notably, the established Brugada-associated gene *SCN5A* (Doundoulakis et al., 2024) did not reach genome-wide significance within this analysis, potentially reflecting limited statistical power, allele frequency differences, or the contribution of alternative loci within this cohort.

#### Takotsubo Cardiomyopathy

For takotsubo cardiomyopathy, two genome-wide significant variants were identified, mapping to *ANKRD31* and *PREX1*. *ANKRD31* encodes a protein containing ankyrin repeat domains involved in protein–protein interactions, while *PREX1* encodes a guanine nucleotide exchange factor involved in RAC1 signalling pathways (NCBI Gene, 2026a, 2026b).

#### Pulmonary Arterial Hypertension

For primary pulmonary arterial hypertension, two genome-wide significant variants were identified, mapping to *ANO10* and *SHF*. *ANO10* belongs to the anoctamin family of calcium-activated chloride channels, while *SHF* is predicted to participate in phosphotyrosine binding and apoptotic signalling pathways (GeneCards, 2026a, 2026b). Dysregulation of apoptosis has previously been implicated in the pathogenesis of pulmonary arterial hypertension (Humbert et al., 2019). ANO10 has been associated with neurological disorders, particularly autosomal recessive cerebellar ataxia. (*MalaCardsSCAR2026*, n.d.; MalaCards, 2026) In addition, infectious and inflammatory signalling pathways, including those related to SARS-CoV-2 infection, have been implicated in the regulation and function of ANO10. (GeneCards, 2026a)

### 3.2 MR identifies LPA as key target with established proteins identified under weaker significance threshold

Only one protein-phenotype relationship remained significant, *LPA* for takotsubo cardiomyopathy, following GSMR analysis. However, several additional protein-phenotype relationships demonstrated suggestive evidence of association and narrowly failed to meet the predefined significance threshold. To better characterise the immediate biological landscape surrounding these rare cardiovascular phenotypes, proteins achieving a weaker significance threshold of *p <* 5*×*10*^−^*^3^ were examined further including CSF3R and APOH for pulmonary arterial hypertension, and ZBTB16 for Takotsubo Cardiomyopathy.

For the Brugada syndrome phenotype, *ENPP2* emerged as a notable candidate. *ENPP2* encodes the principal enzyme (autotaxin) responsible for the production of lysophosphatidic acid, a bioactive lipid mediator implicated in myocardial remodelling, fibrosis, inflammatory signalling, and altered cellular electrophysiology. Dysregulation of these processes may contribute to the development of an arrhythmogenic substrate through structural and functional changes within the myocardium. Additionally, *TNXB* was identified as a candidate protein. *TNXB* encodes tenascin-X, an extracellular matrix glycoprotein involved in maintaining tissue architecture and matrix organisation. Alterations in extracellular matrix composition have been linked to myocardial fibrosis and structural remodelling, both of which can influence conduction velocity, increase conduction heterogeneity, and promote re-entrant electrical circuits.

Collectively, these findings are consistent with the genome-wide association results, which highlighted *THSD7B* and *TRPC4* rather than the canonical *SCN5A* locus. Taken together, the evidence may suggest that structural remodelling, extracellular matrix organisation, and cellular signalling pathways contribute to arrhythmogenic susceptibility alongside established ion-channel mechanisms. This aligns with emerging evidence that Brugada syndrome may represent a more complex disorder involving both electrical and structural myocardial abnormalities rather than a pure channelopathy.

## 4 Discussion

In this study, we performed genome-wide association analyses across three rare cardiovascular diseases and identified both distinct and overlapping patterns of genetic association. While Brugada syndrome, takotsubo cardiomyopathy, and primary pulmonary arterial hypertension are clinically and pathophysiologically distinct entities, several of the identified loci converged on broader biological processes involving; electrophysiological regulation, calcium signalling, cellular remodelling, inflammation, and endothelial dysfunction. These findings suggest that the genetic architecture of these three rare cardiovascular diseases may be more interconnected than is currently appreciated and that common molecular mechanisms may contribute to disease susceptibility across traditionally separate clinical phenotypes.

### Electrophysiological Regulation and Calcium Signalling

Within Brugada syndrome, the strongest association signals were observed within *THSD7B* and *TRPC4* proteins. Although the biological role of *THSD7B* remains incompletely characterised, existing evidence suggests involvement in cytoskeletal organisation and intracellular signalling processes. Structural integrity is increasingly recognised as an important determinant of normal cardiac conduction, influencing sodium-channel localisation, membrane stability, and intracellular anchoring (Bhar-Amato, 2010; Zimmer, Haufe, & Blechschmidt, 2015; Maltsev & Undrovinas, 1997). The identification of *THSD7B* therefore raises the possibility that structural regulatory mechanisms may contribute to arrhythmogenic susceptibility alongside the ion-channel abnormalities traditionally associated with Brugada syndrome.

Additionally, *TRPC4*, a member of the transient receptor potential channel family, is involved in calcium and cation flux regulation and has been implicated in cellular remodelling and arrhythmogenesis (Rios, Sarafian, Camargo, Montezano, & Touyz, 2023; Khan et al., 2024). Network analyses identified interactions between *TRPC4* and *BMPR2*, Dysregulation of BMP-related signalling has previously been associated with altered cardiac development Molecular mechanisms and therapeutic developments of BMPR2 in pulmonary arterial hypertension - ScienceDirect (Jamali, Karamboulas, Rogerson, & Skerjanc, 2001; Wang, Liu, Yang, & Zhou, 2024; Briggs et al., 2008).

Beyond Brugada syndrome, evidence for calcium-dependent mechanisms also emerged within pulmonary arterial hypertension through variants mapping to *ANO10*, a member of the anoctamin family of calcium-activated chloride channels. Although the cardiovascular role of *ANO10* remains poorly characterised, these observations collectively support a broader role for calcium signalling abnormalities across multiple rare cardiovascular phenotypes.

### Cellular Remodelling

A second recurring theme was the involvement of pathways related to tissue/cellular remodelling, fibrosis, apoptosis, and proliferative regulation. *ENPP2* was seen to be just below significance level for Brugada syndrome, however it known to encode autotaxin, the enzyme responsible for generating lysophosphatidic acid. Lysophosphatidic acid is a potent signalling molecule involved in inflammation, fibrosis, and myocardial remodelling (Cholia, Nayyar, Kumar, & Mantha, 2015; F. Huang et al., 2026). Structural abnormalities and localised fibrosis of the right ventricular outflow tract have increasingly been recognised in Brugada syndrome and are thought to contribute to arrhythmogenesis (Nademanee et al., 2015).

Evidence for remodelling-related mechanisms was also observed in pulmonary arterial hypertension. Variants mapping to *SHF* suggest potential involvement of signalling pathways governing apoptosis and cellular proliferation (Paulin, Meloche, & Bonnet, 2012; Humbert et al., 2019). Excessive smooth-muscle proliferation, resistance to apoptosis, and progressive vascular restructuring are hallmarks of primary pulmonary arterial hypertension pathogenesis (Thenappan, Ormiston, Ryan, & Archer, 2018), making these findings biologically plausible within the context of current disease models. Taken together, the emergence of loci linked to fibrosis, apoptosis, and proliferative regulation across multiple phenotypes suggests that dysregulated cellular/tissue remodelling may represent a common contributor to rare cardiovascular disease susceptibility despite differing clinical manifestations.

### Inflammatory and Endothelial Pathways

Several loci associated with pulmonary arterial hypertension approached genome-wide significance, including *CSF3R* and *APOH*. *CSF3R* participates in inflammatory signalling and may influence disease susceptibility through effects on immune activation and endothelial remodelling (Dong et al., 2021). Likewise, *APOH* encodes *β*2-glycoprotein I, a multifunctional protein involved in immune-complex interactions, coagulation, and endothelial regulation. (Castro et al., 2010). Although neither locus reached genome-wide significance, both are consistent with growing evidence that chronic inflammation and endothelial dysfunction play central roles in primary pulmonary arterial hypertension development and progression.

Further support for endothelial mechanisms emerged through the identification of *ZBTB16*, with associations observed just below statistical thresholds for both takotsubo cardiomyopathy and pulmonary arterial hypertension analyses. *ZBTB16* functions as a transcriptional regulator involved in inflammatory responses, vascular homeostasis, and cellular differentiation (Tavosanis, 2026). Impaired microvascular resilience and endothelial dysfunction have long been proposed as important contributors to takotsubo cardiomyopathy, particularly in the context of catecholamine-mediated myocardial injury. (Yosofi et al., 2026) The appearance of *ZBTB16* across multiple phenotypes therefore raises the possibility that pathways governing vascular repair and endothelial integrity may represent shared determinants of cardiovascular vulnerability.

### Limitations and Conclusions

Several limitations should be considered when interpreting these findings. Although UK Biobank represents one of the largest population-based resources currently available, the rarity of the investigated phenotypes resulted in relatively modest case numbers. Consequently, statistical power remained limited for the detection of variants with small effect sizes and may have reduced our ability to identify associations within established disease loci. This limitation may partially explain the absence of genome-wide significant associations within genes such as *SCN5A*, despite their recognised role in Brugada syndrome. The phenotypes were derived primarily from hospital episode statistics and ICD-based coding. While this approach enables investigation of rare diseases at a scale that would otherwise be difficult to achieve, diagnostic coding is inherently imperfect and may introduce phenotypic heterogeneity. Conditions such as Brugada syndrome and takotsubo cardiomyopathy often require specialist clinical investigations for definitive diagnosis and may therefore be susceptible to underdiagnosis or misclassification within routine healthcare records. Any resulting non-differential misclassification would be expected to attenuate true genetic associations and reduce statistical power.

To minimise population stratification, analyses were restricted to individuals of European genetic ancestry. While this improves internal validity, it limits the generalisability of the findings to other populations. Differences in allele frequencies, linkage disequilibrium structure, and environmental exposures may influence both the magnitude and detectability of genetic associations across ancestries. Validation in more diverse cohorts will therefore be important to establish the broader relevance of the identified loci. An additional limitation is the absence of an independent replication cohort. Replication remains a cornerstone of genetic discovery, particularly for studies of rare diseases where statistical power is inherently constrained. Although several associations achieved conventional genome-wide significance thresholds, confirmation within independent datasets will be required before firm conclusions regarding novel disease loci can be drawn. Finally, while several biologically plausible mechanisms were proposed, genome-wide association studies identify statistical associations rather than causal pathways. Functional studies will therefore be necessary to determine how the identified variants influence cardiovascular biology and whether they contribute directly to disease pathogenesis.

These findings reinforce the substantial genetic heterogeneity underlying rare cardiovascular diseases while simultaneously highlighting common biological themes involving electrophysiological regulation, calcium signalling, cellular remodelling, inflammation, and endothelial dysfunction. Rather than representing entirely distinct pathological entities, these conditions may share elements of a broader molecular framework that contributes to disease susceptibility through different downstream manifestations. Integrating genomic association analyses with functional annotation, proteomic data, and causal inference approaches may therefore provide valuable opportunities for identifying biologically relevant pathways and prioritising therapeutic targets within rare cardiovascular disease research.

## Data Availability

All data produced in the present study are available upon reasonable request to the authors

## A Non-retained mapping SNPs

rs11211569, rs141369406, rs10948570, rs10948570, rs13213461, rs550271061, rs28605277, rs10009814, rs12331976, rs7695743, rs74387669, rs76831512, rs9995940, rs28587453, rs112312121, rs112728295.

## Supplementary Figures

**Figure 4:**
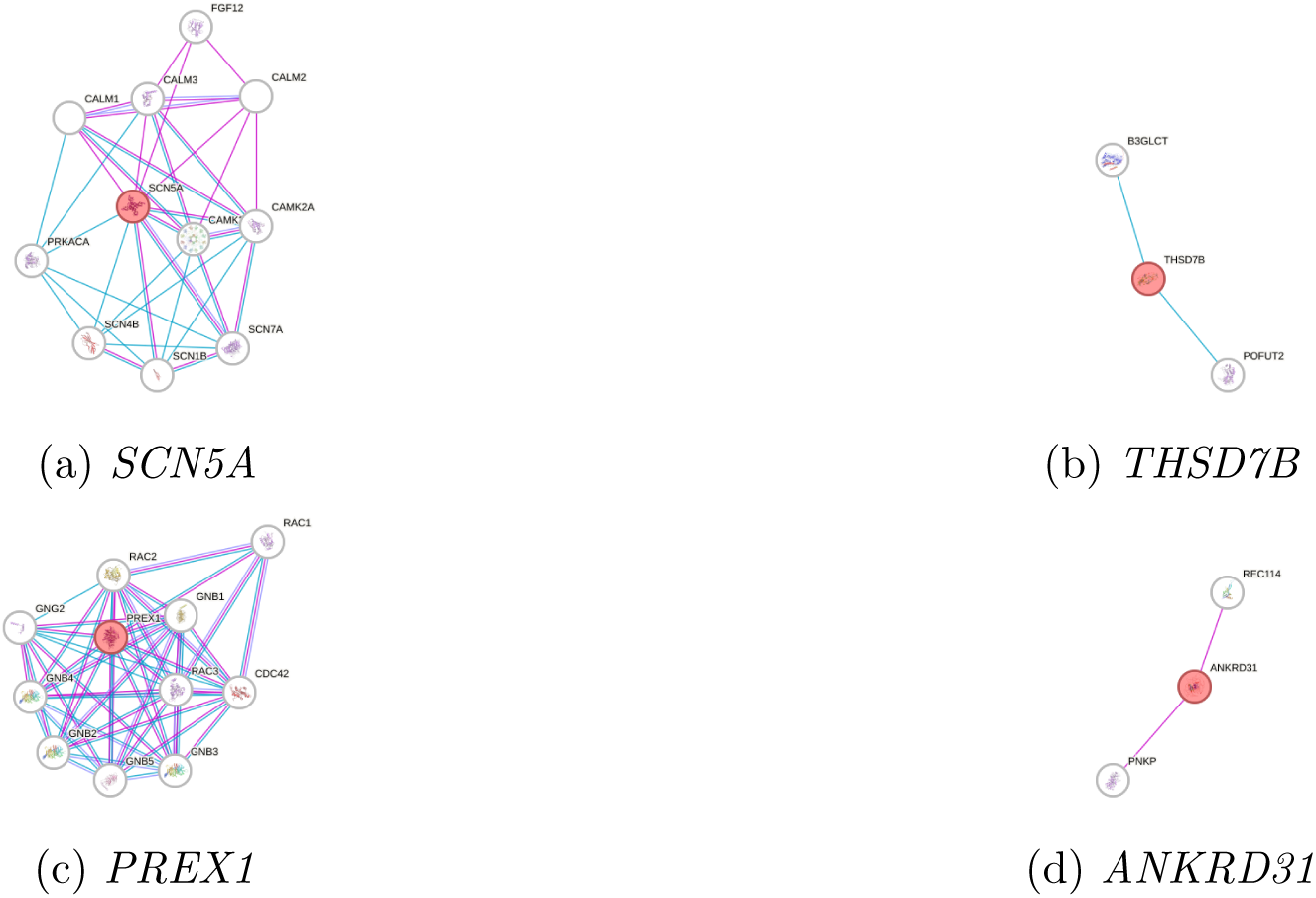
STRING protein–protein interaction networks for additional loci identified through genome-wide association analyses. Nodes represent proteins and edges represent known or predicted interactions derived from curated databases and experimentally validated evidence.

## References

(n.d.).

Antzelevitch, C. (2006). Brugada syndrome. Pacing and Clinical Electrophysiology, 29 (10), 1130–1159. doi: 10.1111/j.1540-8159.2006.00507.x

Bhar-Amato, N. L.. L. P., J. (2010). A review of the mechanisms of ventricular arrhythmia in brugada syndrome. Indian pacing and electrophysiology journal, 10 (9), 410–425.

Borchard, U., & Hafner, D. (2000). Ion channels and arrhythmias. Zeitschrift für Kardiologie, 89 (Suppl 3), 6–12. (Article in German)

Briggs, L. E., Takeda, M., Cuadra, A. E., Wakimoto, H., Marks, M. H., Walker, A. J., . . . Kasahara, H. (2008). Perinatal loss of nkx2-5 results in rapid conduction and contraction defects. Circulation Research, 103 (6), 580–590. Retrieved from https://www.ncbi.nlm.nih.gov/pmc/articles/PMC2590500/ doi: 10.1161/CIRCRESAHA.108.171835

British Heart Foundation. (2026). Takotsubo cardiomyopathy (broken heart syndrome). Retrieved 2026-07-03, from https://www.bhf.org.uk/informationsupport/conditions/takotsubo-cardiomyopathy

Castro, A., Lázaro, I., Selva, D. M., Céspedes, E., Girona, J., Plana, N., . . . Masana, L. (2010, March). Apoh is increased in the plasma and liver of type 2 diabetic patients with metabolic syndrome. Atherosclerosis.

Cholia, R. P., Nayyar, H., Kumar, R., & Mantha, A. K. (2015). Understanding the multifaceted role of ectonucleotide pyrophosphatase/phosphodiesterase 2 (enpp2) and its altered behaviour in human diseases. Current Molecular Medicine, 15 (10), 932–943. doi: 10.2174/1566524015666150921104804

Dong, H., Li, X., Cai, M., Zhang, C., Mao, W., Wang, Y., . . . Huang, X. (2021). Integrated bioinformatic analysis reveals the underlying molecular mechanism of and potential drugs for pulmonary arterial hypertension. Aging (Albany NY), 13 (10), 14234–14257. doi: 10.18632/aging.203040

Doundoulakis, I., Pannone, L., Chiotis, S., Della Rocca, D. G., Sorgente, A., Tsioufis, P., . . . de Asmundis, C. (2024). Scn5a gene variants and arrhythmic risk in brugada syndrome: An updated systematic review and meta-analysis. Heart Rhythm, 21 (10), 1987–1997. Retrieved from https://www.sciencedirect.com/science/article/pii/S1547527124023749 (Focus Issue: Sudden Death) doi: 10.1016/j.hrthm.2024.04.047

Frak, W., Wojtasińska, A., Lisińska, W., M-lynarska, E., Franczyk, B., & Rysz, J. (2022). Pathophysiology of cardiovascular diseases: New insights into molecular mechanisms of atherosclerosis, arterial hypertension, and coronary artery disease. Biomedicines, 10 (8). Retrieved from https://www.mdpi.com/2227-9059/10/8/1938 doi: 10.3390/biomedicines10081938

GeneCards. (2025). Genecards: The human gene database. https://www.genecards.org/. (Last Updated November 10, 2025)

GeneCards. (2026a). Ano10 gene - genecards human gene database. https://www.genecards.org/cgi-bin/carddisp.pl?gene=ANO10. (Accessed Feb 2026)

GeneCards. (2026b). Shf gene - genecards human gene database. https://www.genecards.org/cgi-bin/carddisp.pl?gene=SHF. (Accessed Feb 2026)

Ghadri, J.-R., Wittstein, I. S., Prasad, A., Sharkey, S., Dote, K., Akashi, Y. J., . . . Templin, C. (2018, 06). International expert consensus document on takotsubo syndrome (part i): Clinical characteristics, diagnostic criteria, and pathophysiology. European Heart Journal, 39 (22), 2032–2046. Retrieved from 10.1093/eurheartj/ehy076 doi: 10.1093/eurheartj/ehy076

Great Ormond Street Hospital. (2015). Brugada syndrome. Retrieved from https://www.gosh.nhs.uk/conditions-and-treatments/conditions-we-treat/brugada-syndrome/ (Last reviewed: December 2015. Accessed: 2026-04-14)

Hoendermis, E. S. (2011). Pulmonary arterial hypertension: an update. Netherlands Heart Journal, 19 (12), 514–522. doi: 10.1007/s12471-011-0222-1

Huang, F., Zhou, M., Chen, Y., Hua, S., Han, Y., Fan, Y., . . . Jin, W. (2026). Cross-dataset transcriptomic analyses identify a conserved enpp2+ macrophage-fibroblast activation axis in hypertrophic cardiomyopathy. Briefings in Bioinformatics, 27 (1), bbag036. Retrieved from 10.1093/bib/bbag036 doi: 10.1093/bib/bbag036

Huang, H., Hu, C., Zhang, R., Xu, H., Cao, M., & Fu, Y. (2025). Global burden of pulmonary arterial hypertension and associated heart failure. JACC: Heart Failure, 13 (10), 102385. Retrieved from https://www.jacc.org/doi/abs/10.1016/j.jchf.2024.12.005 doi: 10.1016/j.jchf.2024.12.005

Humbert, M., Guignabert, C., Bonnet, S., Dorfmüller, P., Klinger, J. R., Nicolls, M. R.,. . . Rabinovitch, M. (2019). Pathology and pathobiology of pulmonary hypertension: state of the art and research perspectives. European Respiratory Journal, 53 (1). Retrieved from https://publications.ersnet.org//content/erj/53/1/1801887 doi: 10.1183/13993003.01887-2018

Jamali, M., Karamboulas, C., Rogerson, P. J., & Skerjanc, I. S. (2001). Bmp signaling regulates nkx2-5 activity during cardiomyogenesis. FEBS Letters, 509 (1), 126–130. Retrieved from https://pubmed.ncbi.nlm.nih.gov/11734219/ doi: 10.1016/S0014-5793(01)03151-9

Khan, S. U., Khan, S. U., Suleman, M., Khan, M. U., Alsuhaibani, A. M., Refat, M. S., . . . Saeed, S. (2024). The multifunctional trpc6 protein: Significance in the field of cardiovascular studies. Current Problems in Cardiology, 49 (1, Part B), 102112. Retrieved from https://www.sciencedirect.com/science/article/pii/S0146280623005297 doi: 10.1016/j.cpcardiol.2023.102112

Lai, Y.-C., Potoka, K. C., Champion, H. C., Mora, A. L., & Gladwin, M. T. (2014). Pulmonary arterial hypertension: The clinical syndrome. Circulation Research, 115 (1), 115–130. doi: 10.1161/CIRCRESAHA.115.301146

Lan, N. S. H., Massam, B. D., Kulkarni, S. S., & Lang, C. C. (2018). Pulmonary arterial hypertension: Pathophysiology and treatment. Diseases, 6 (2), 38. doi: 10.3390/diseases6020038

Li, C., Xu, K., Du, A., Fu, N., Xu, Z., & Chang, Q. (2025). Global, regional and national epidemiology of myocarditis: health inequalities, risk factors and forecasted burden based on the global burden of disease study 2021. Heart, 111 (18), 867– 876. Retrieved from https://heart.bmj.com/content/111/18/867 doi: 10.1136/heartjnl-2024-325523

MalaCards. (2026). Autosomal recessive cerebellar ataxia (arca). Retrieved from https://www.malacards.org/card/autosomal_recessive_cerebellar_ataxia (Accessed: 2026-04-14)

Maltsev, V. A., & Undrovinas, A. I. (1997). Cytoskeleton modulates coupling between availability and activation of cardiac sodium channel. American Journal of Physiology-Heart and Circulatory Physiology , 273 (4), H1832–H1840. Retrieved from 10.1152/ajpheart.1997.273.4.H1832 (PMID: 9362250) doi: 10.1152/ajpheart.1997.273.4.H1832

Minhas, A. S., &, et al. (2015). Nationwide trends in reported incidence of takotsubo cardiomyopathy. The American Journal of Cardiology , 116 (10), 1584–1590. doi: 10.1016/S0002-9149(15)01633-1

Mizusawa, Y., & Wilde, A. A. M. (2012). Brugada syndrome. Circulation: Arrhythmia and Electrophysiology , 5 (3), 606–616. doi: 10.1161/CIRCEP.111.964577

Nademanee, K., Raju, H., de Noronha, S. V., Papadakis, M., Robinson, L., Rothery, S., . . . Behr, E. R. (2015). Fibrosis, connexin-43, and conduction abnormalities in the brugada syndrome. Journal of the American College of Cardiology , 66 (18), 1976–1986. doi: 10.1016/j.jacc.2015.08.862

National Institute for Health and Care Research. (2025). New £50m funding call to tackle inequalities in cardiovascular disease. https://www.nihr.ac.uk/news/new-50m-funding-to-tackle-inequalities-cardiovascular-disease. (Accessed: 2025-12-01)

NCBI Gene. (2026a). Ankrd31 ankyrin repeat domain 31 [homo sapiens]. https://www.ncbi.nlm.nih.gov/gene/256006. (Accessed Feb 2026)

NCBI Gene. (2026b). Prex1 phosphatidylinositol-3,4,5-trisphosphate dependent rac exchange factor 1 [homo sapiens]. https://www.ncbi.nlm.nih.gov/gene/57580. (Accessed Feb 2026)

NHS. (2026). Brugada syndrome. https://www.nhs.uk/conditions/brugada-syndrome/. (Accessed: 2026-07-03)

Paulin, R., Meloche, J., & Bonnet, S. (2012). Stat3 signaling in pulmonary arterial hypertension. JAK-STAT , 1 (4), 223–233. Retrieved from https://pubmed.ncbi.nlm.nih.gov/24058777/ (Accessed: 2026-05-07) doi: 10.4161/jkst.22366

Pelliccia, F., Kaski, J. C., Crea, F., & Camici, P. G. (2017). Pathophysiology of takotsubo syndrome. Circulation, 135 (24), 2426–2441. doi: 10.1161/CIRCULATIONAHA.116.027121

Rattanawong, P., et al. (2025). Prevalence and incidence of type 1 brugada pattern at mayo clinic during 30 years. Mayo Clin. Proc.. Online ahead of print. Retrieved from https://www.mayoclinicproceedings.org/article/S0025-6196(24)00303-3/abstract (S0025-6196(24)00303-3) doi: 10.1016/j.mayocp.2024.???.S0025-6196(24)00303-3

Rios, F. J., Sarafian, R. D., Camargo, L. L., Montezano, A. C., & Touyz, R. M. (2023). Recent advances in understanding the mechanistic role of transient receptor potential ion channels in patients with hypertension. Canadian Journal of Cardiology , 39 (12), 1859–1873. Retrieved from https://www.sciencedirect.com/science/article/pii/S0828282X23018378 (Focus Issue: Novel Molecular Mechanisms and Therapeutics in Hypertension and Vascular Prevention: Genetics and Beyond) doi: 10.1016/j.cjca.2023.10.009

Roth, G. A., Mensah, G. A., Johnson, C. O., Addolorato, G., Ammirati, E., Baddour, L. M., . . . Fuster, V. (2020). Global burden of cardiovascular diseases and risk factors, 1990–2019: Update from the gbd 2019 study. Journal of the American College of Cardiology , 76 (25), 2982–3021. Retrieved from https://www.sciencedirect.com/science/article/pii/S0735109720377755 doi: 10.1016/j.jacc.2020.11.010

Scafa-Udriste, A., Horodinschi, R. N., Babos, M., & Dinu, B. (2024). Diagnostic challenges between takotsubo cardiomyopathy and acute myocardial infarction—where is the emergency?: a literature review. International Journal of Emergency Medicine, 17 , 22. Retrieved from https://www.ncbi.nlm.nih.gov/pmc/articles/PMC10870686/ doi: 10.1186/s12245-024-00544-3

Shukla, K., Basile, E. J., Van Name, J. P., & Ahmed, I. (2025). Brugada syndrome. Treasure Island (FL): StatPearls Publishing. Retrieved from https://www.ncbi.nlm.nih.gov/books/NBK519568/ (Updated December 1, 2025)

Sun, B. B., Maranville, J. C., Peters, J. E., Stacey, D., Staley, J. R., Blackshaw, J., . . . Butterworth, A. S. (2018). Genomic atlas of the human plasma proteome. Nature, 558 (7708), 73–79. Retrieved from 10.1038/s41586-018-0175-2 doi: 10.1038/s41586-018-0175-2

Tavosanis, A. (2026). Endothelial zbtb16 preserves vascular health and cardiac function during aging. Nature Cardiovascular Research. Retrieved from 10.1038/s44161-026-00781-y (Research Highlight, published 2026-02-02) doi: 10.1038/s44161-026-00781-y

Thenappan, T., Ormiston, M. L., Ryan, J. J., & Archer, S. L. (2018). Pulmonary arterial hypertension: pathogenesis and clinical management. BMJ , 360 , j5492. doi: 10.1136/bmj.j5492

UK Parliament. (2025). Written question 26140. Retrieved 2025-12-01, from https://questions-statements.parliament.uk/written-questions/detail/2025-01-23/26140

Wang, Z., Liu, X.-Y., Yang, C.-X., & Zhou, H.-M. (2024). Discovery and functional investigation of bmp4 as a new causative gene for human congenital heart disease. Disease Markers, 2024 , 2034–2048. Retrieved from https://www.ncbi.nlm.nih.gov/pmc/articles/PMC11170606/ doi: 10.62347/DGCD4269

WHO. (2025). Cardiovascular diseases (cvds). Retrieved 2026-02-16, from https://www.who.int/news-room/fact-sheets/detail/cardiovascular-diseases-(cvds)

Yosofi, B., de Waard, G., van de Hoef, T. P., Maas, A. H. E. M., Khan, H., Dawson, D., . . . Damman, P. (2026). Role of coronary microvascular dysfunction in takotsubo syndrome. Heart , heartjnl-2025–326929. (Online ahead of print) doi: 10.1136/heartjnl-2025-326929

Zimmer, T., Haufe, V., & Blechschmidt, S. (2015, March). Voltage-gated sodium channels in the mammalian heart. Global Cardiology Science and Practice, 2014 (4). Retrieved from 10.5339/gcsp.2014.58 doi: 10.5339/gcsp.2014.58

